# The Value of the Gensini Score For Prognostic Assessment in Patients with Acute Coronary Syndrome--A Retrospective Cohort Study Based on Machine Learning Methods

**DOI:** 10.1101/2023.09.06.23295161

**Authors:** Lixia Chen, Sixiang Jia, Xuanting Mou, Yiting Tu, Wenting Lin, Feng Chao, Shudong Xia

## Abstract

**Background:** The Gensini score (GS) provides a good assessment of the degree of coronary plate loading. However, its clinical significance has been little explored.

**Methods:** In this retrospective cohort study, we implemented model development and performance comparison on database of The Fourth Affiliated Hospital of Zhejiang University School of Medicine (2019.1-2020.12). The patients were followed up for 2 years. Follow-up endpoint was the occurrence of MACCEs. We extracted clinical baseline data from each ACS patient within 24 hours of hospital admission and randomly divided the datasets into 70% for model training and 30% for model validation. Area under the curve (AUC) was used to compare the prediction performance of XGBoost, SGD and KNN. A decision tree model was constructed to predict the probability of MACCEs using a combination of weight features picked by XGBoost and clinical significance.

**Results:** A total of 361 ACS patients who met the study criteria were included in this study. It could be observed that the probability of a recurrent MACCEs within 2 years was 25.2%. XGboost had the best predictive efficacy (AUC:0.97). GS has high clinical significance. Then we used GS, Age and CK-MB to construct a decision tree model to predict the probability model of MACCEs reoccurring, and the final AUC value reached 0.771.

**Conclusions:** GS is a powerful indicator for assessing the prognosis of patients with ACS. The cut-off value of GS in the decision tree model provides a reference standard for grading the risk level of patients with ACS.

## 1. Introduction

Currently, the incidence of Acute Coronary Syndrome (ACS) remains high globally. As an acute episode of coronary heart disease (CAD), ACS has a relatively higher mortality and morbidity [1,2]. The Framingham Heart Study’s 10-year follow-up data revealed that the incidence of ACS events increases dramatically with age [3,4].

For ACS, percutaneous coronary intervention (PCI) has been formalized as an effective treatment [5,6]. Patients with ACS often have a combination of clinical risk factors, such as hypertension, cerebral infarction, diabetes mellitus, smoking, and so on. Therefore they have a higher probability of recurrent Major Adverse Cardiac and Cerebrovascular Events (MACCEs) after emergency PCI [7,8].

Scholars at home and abroad have launched a large number of studies to investigate the prediction of long-term MACCEs in patients with ACS. They developed indicators like TACE/ADAM17, Triglyceride-glucose index, HEART SCORE, Systemic immune-inflammation index (SII) and so on [7,9-11]. These predictors have achieved relatively satisfactory results. Unfortunately, none of them are directly linked to the ACS, but rather an indirectly mediated.

The Gensini score (GS) provides an objective assessment of the degree of coronary plaque load [12,13]. Wang KY et al. study has revealed the predictive value of GS in patients with CAD undergoing PCI [14]. However, they have not yet developed a valid GS-based predictive model for patients with CAD. Nowadays, Machine Learning (ML) models based on Artificial Intelligence (AI) theory are relatively mature in medicine [15,16]. ML has the advantage of being good at extracting non-linear relationships between different variables and showing researchers the weighting relationships by feature maps.

Adoption of the above background, we propose to apply ML approaches to elucidate the prognostic value of GS for ACS patients. Further more, to construct a GS-based prediction model the probability of a recurrent MACCEs within 2 years for ACS patients.

## 2. Methods

### 2.1 Date Source

All patients in the database of the Chest Pain Center of the Fourth Affiliated Hospital of Zhejiang University School of Medicine who were seen between 2019.1-2020.12 were included.

### 2.2 Ethical Considerations

The Ethics Committee of the Fourth Affiliated Hospital of Zhejiang University School of Medicine approved the study. The study adhered to the Declaration of Helsinki.

### 2.3 Study population

A total of 455 patients who needed medical attention for acute chest pain were exported from the database.The exclusion criteria for this study were as follows:

1. the patient has already received a previous coronary intervention
2. Suspected type II myocardial infarction [17,18]
3. Chest pain of non-cardiac origin [19]
4. Seriously missing basic data
5. Abandonment of interventional treatment for their own reasons, opting for conservative treatment
6. Acute respiratory and cardiac arrest leading to failure of resuscitation

In this retrospective study, we examined whether MACCEs occurred within 2 years as the primary outcome. The follow-up date was up to 2023.2. We regularly followed up patients with ACS who underwent PCI at 1, 3, 6, 12, and 24 months, and if MACCEs occurred during this period, the follow-up was stopped immediately. At the duration of follow-up, the patient’s clinical information stored in the electronic medical record system was again evaluated and confirmed, including the patient’s previous underlying diseases (Hypertension, Diabetes Mellitus, Coronary Artery Disease,etc.), as well as the patient’s current clinical symptoms. The emergency coronary angiography and coronary stent implantation procedures were performed by 2 experienced coronary interventionists in our cardiology.

Routine hematology, liver and renal function, cardiac enzyme profile, NT-proBNP and troponin levels were recorded within 24 hours of the patient’s initial admission, and all of the above were available from our laboratory. All follow-up information was available in the Chest Pain Center database, and data that were not available were supplemented by telephone contact.

A total of 361 patients ultimately met the criteria for this study.

### 2.4 Gensini Score

For patients who met the study criteria, we performed GS calculations based on coronary angiography results. The degree of stenosis of coronary lesions was assessed using the coronary Gensini Score as follows: first, the largest stenotic lesions in each branch artery were quantitatively assessed on the basis of coronary angiography, and the degree of stenosis ≤25%, 26% to 50%, 51% to 75%, 76% to 90%, 91% to 99%, and 100% were recorded as 1, 2, 4, 8, 16 and 32 points, respectively. Then, according to the different coronary branches, the above scores were multiplied by the corresponding values:① Left Main artery (LM)×5.0. ② Left anterior descending branch (LAD): proximal LAD×2.5, middle LAD×1.5, distal LAD×1.0. ③ First diagonal branch (D1)×1.0, second diagonal branch (D2)×0.5. ④ Left levator branch (LCX): proximal LCX×2.5, middle LCX×1.5, middle LCX×1.5, and distal LCX×1.0. ⑤ Obtuse marginal branch (OM)×1.0. ⑥ Left posterior lateral branch (PL)×0.5. ⑦ Right coronary artery (RCA)×1.0. ⑧ Posterior descending branch (PDA)×1.0. Finally, the total score of each lesion was summed to obtain the patient’s coronary Gensini score. In this study, the degree of coronary stenosis evaluated with GS was classified on a 4-point scale: none (GS=0), mild (0 GS≥20), moderate (20≤GS<50) or severe (GS≥50).

### 2.5 Statistical analysis

All statistical analysis and calculations were performed using R software (version 4.3.1) and Python (version 3.9.0). Categorical variables are expressed as total numbers and percentages. Continuous variables are expressed as mean±standard deviation. *P* < 0.05 was considered statistically significant for this study.

Given the multidimensional characteristics among the variables, we first used Pearson’s heat map to observe the correlation among the variables and used the t-test method for data downscaling [20]. Three ML models -XGBoost, SGD, and KNN were used to develop predictive models [21-25]. The downscaled data is fed into a subsequent ML model. The predictive performance of each model was evaluated by the AUC. In addition, we calculated the Recall, Positive Predictive Value (PPV), Negative Predictive Value (NPV), and F1 Score. In addition, to evaluate the utility of the decision models by quantifying the net benefit at different threshold probabilities, we performed Decision Curve Analysis (DCA) [26,27]. Considering the single-center data, we also used K-fold cross-validation to check the stability of our model.

We use the optimal predictive model to observe the weighting characteristics of the variables. Selected features were combined with clinical significance to construct a Decision Tree model to predict the incidence of MACCEs in patients over a 2-year period [28]. And its decision-making performance is evaluated by AUC. If necessary, we will focus on the prognosis of this indicator using survival analysis curves.

The flow of this study is shown schematically in **Figure 1**.

**Figure 1.**
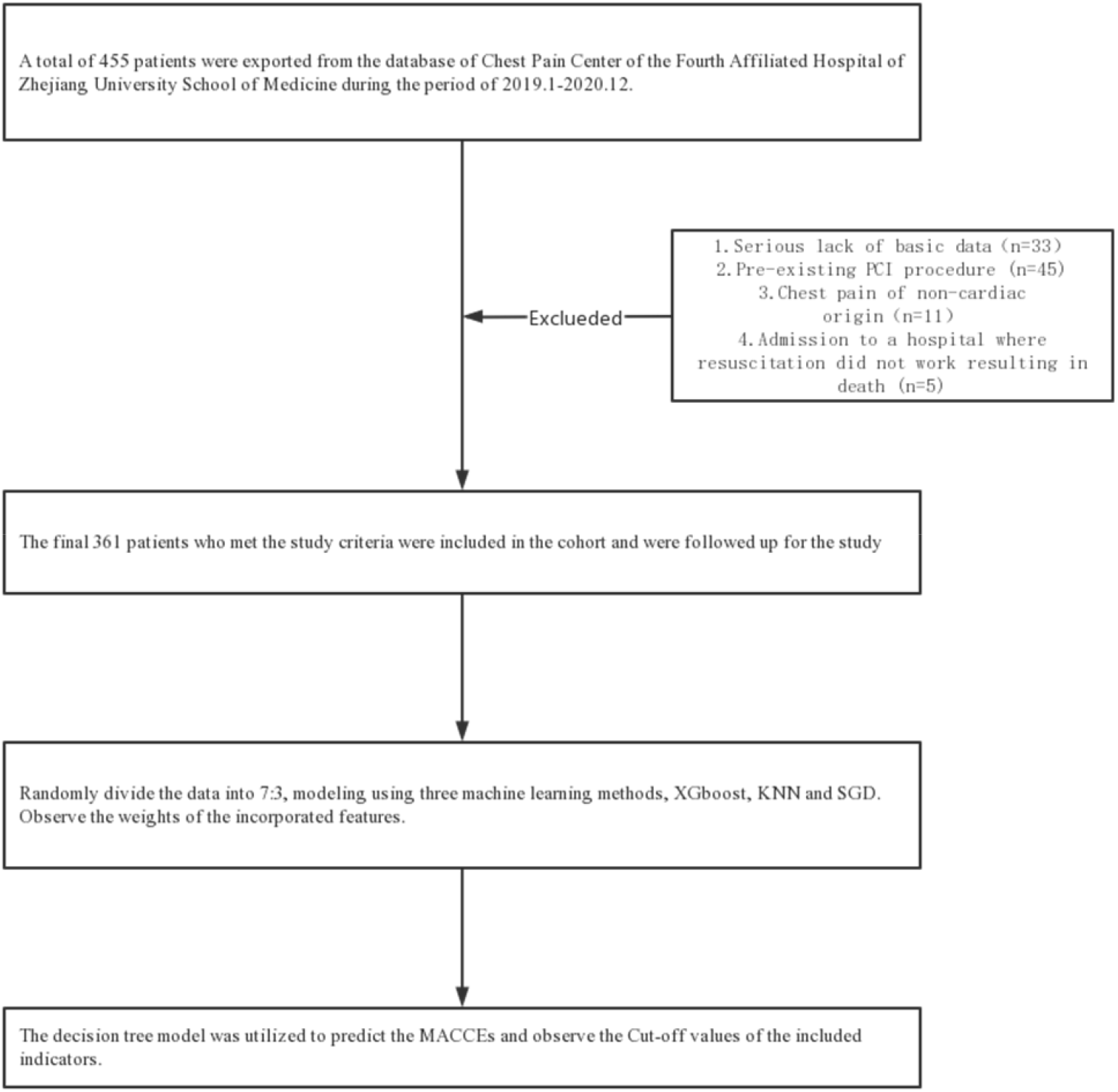
Schematic diagram of the flow of this study.

## 3. Results

### 3.1 Patient Characteristics

As seen in Table 1, a total of 361 patients were included in the study. 91 patients (25.2%) had MACCEs events within 2 years. Among the Table 1, GS showed a statistically significant difference within the two groups mentioned above (*P*<0.001). To facilitate machine learning, we set the time-years in which MACCEs occur as independent labels as the annotation of the data.

**Table 1.** Characteristics of subjects included in the study.

| Baseline information on subjects |  |  |  |  |  |
| --- | --- | --- | --- | --- | --- |
|  |  | Overall | None | MACCEs | P-Value |
| N |  | 361 | 270 | 91 |  |
| Sex, n (%) | Male | 76 (21.1) | 59 (21.9) | 17 (18.7) | 0.622 |
|  | Female | 285 (78.9) | 211 (78.1) | 74 (81.3) |  |
| Age, mean (SD) |  | 63.1 | 61.9 | 66.7 | 0.005 |
|  |  | (14.4) | (14.5) | (13.5) |  |
| GS, mean (SD) |  | 63.6 | 59.0 | 77.1 | <0.001 |
|  |  | (40.6) | (39.4) | (41.2) |  |
| CK, mean (SD) |  | 347.7 | 339.7 | 371.4 | 0.626 |
|  |  | (561.4) | (574.7) | (522.1) |  |
| CK-MB, mean (SD) |  | 54.3 | 56.2 | 48.8 | 0.697 |
|  |  | (251.1) | (288.0) | (65.1) |  |
| LDH, mean (SD) |  | 289.1 | 291.2 | 282.6 | 0.778 |
|  |  | (367.4) | (414.5) | (162.7) |  |
| ALT, mean (SD) |  | 59.1 | 59.8 | 56.8 | 0.763 |
|  |  | (113.8) | (127.2) | (58.9) |  |
| Troponin, mean (SD) |  | 1.0 (3.7) | 1.2 (4.2) | 0.5 (1.1) | 0.017 |
| PT, mean (SD) |  | 10.7 (5.6) | 10.8 (6.3) | 10.5 (1.9) | 0.572 |
| INR, mean (SD) |  | 2.3 (21.2) | 2.7 (24.6) | 1.0 (0.2) | 0.233 |
| APTT, mean (SD) |  | 35.0 | 37.6 | 27.1 (8.2) | 0.274 |
|  |  | (136.3) | (157.5) |  |  |
| TT, mean (SD) |  | 20.0 | 20.9 | 17.3 (3.2) | 0.008 |
|  |  | (18.9) | (21.7) |  |  |
| Fibrinogen, mean (SD) |  | 3.2 (1.4) | 3.2 (1.1) | 3.5 (2.1) | 0.142 |
| Hct, mean (SD) |  | 42.9 (7.3) | 43.0 (6.8) | 42.4 (8.7) | 0.526 |
| CRP, mean (SD) |  | 121.5 | 157.9 | 13.6 | 0.323 |
|  |  | (2072.1) | (2395.9) | (33.6) |  |
| Scr, mean (SD) |  | 91.5 | 93.4 | 85.7 | 0.287 |
|  |  | (89.1) | (100.9) | (36.2) |  |
| K, mean (SD) |  | 3.8 (0.5) | 3.8 (0.5) | 3.8 (0.7) | 0.754 |
| Total cholesterol, mean (SD) |  | 3.5 (1.8) | 3.4 (1.9) | 3.8 (1.4) | 0.019 |
| Triglyceride, mean (SD) |  | 1.6 (2.0) | 1.5 (2.0) | 1.9 (1.9) | 0.083 |
| High-density lipoprotein, mean (SD) |  | 0.9 (0.5) | 0.9 (0.5) | 1.0 (0.4) | 0.065 |
| Low-density lipoprotein, mean (SD) |  | 1.8 (1.1) | 1.8 (1.1) | 2.0 (0.9) | 0.064 |
| Blood platelets, mean (SD) |  | 216.0 | 221.6 | 199.3 | 0.011 |
|  |  | (88.0) | (94.8) | (61.5) |  |
| Neutrophils, mean (SD) |  | 6.8 (3.2) | 6.9 (3.3) | 6.6 (2.7) | 0.294 |
| Lymphocytes, mean (SD) |  | 2.0 (1.3) | 2.0 (1.2) | 2.0 (1.6) | 0.995 |
| SII, mean (SD) |  | 1092.3 | 1147.0 | 930.1 | 0.085 |
|  |  | (1375.1) | (1516.5) | (808.9) |  |
| NT-proBNP, mean (SD) |  | 1466.4 | 1520.2 | 1306.5 | 0.626 |
|  |  | (4237.7) | (4526.4) | (3250.0) |  |
| Diabetes, n (%) | N | 291 (80.6) | 221 (81.9) | 70 (76.9) | 0.381 |
|  | Y | 70 (19.4) | 49 (18.1) | 21 (23.1) |  |
| Hypertension, n (%) | N | 205 (56.8) | 163 (60.4) | 42 (46.2) | 0.025 |
|  | Y | 156 (43.2) | 107 (39.6) | 49 (53.8) |  |
| Stroke, n (%) | N | 339 (93.9) | 253 (93.7) | 86 (94.5) | 0.982 |
|  | Y | 22 (6.1) | 17 (6.3) | 5 (5.5) |  |
| Smoking, n (%) | N | 206 (57.1) | 151 (55.9) | 55 (60.4) | 0.529 |
|  | Y | 155 (42.9) | 119 (44.1) | 36 (39.6) |  |
| NLR, mean (SD) |  | 5.1 (5.5) | 5.2 (5.9) | 4.7 (3.8) | 0.384 |
| PLR, mean (SD) |  | 142.6<br>(108.7) | 146.6<br>(118.1) | 130.7<br>(73.8) | 0.134 |
| Pain Score, mean (SD) |  | 1.1 (1.2) | 1.1 (1.2) | 1.1 (1.2) | 0.976 |
| D-D, mean (SD) |  | 1.7 (8.6) | 1.8 (9.5) | 1.3 (5.4) | 0.507 |
| Expired infarction, n (%) | N | 237 (65.7) | 178 (65.9) | 59 (64.8) | 0.951 |
|  | Y | 124 (34.3) | 92 (34.1) | 32 (35.2) |  |
| One month, n (%) | N | 343 (95.0) | 270<br>(100.0) | 73 (80.2) | <0.001 |
|  | Y | 18 (5.0) |  | 18 (19.8) |  |
| Three months, n (%) | N | 346 (95.8) | 270<br>(100.0) | 76 (83.5) | <0.001 |
|  | Y | 15 (4.2) |  | 15 (16.5) |  |
| Six months, n (%) | N | 355 (98.3) | 270<br>(100.0) | 85 (93.4) | <0.001 |
|  | Y | 6 (1.7) |  | 6 (6.6) |  |
| Twelve months, n (%) | N | 320 (88.6) | 270<br>(100.0) | 50 (54.9) | <0.001 |
|  | Y | 41 (11.4) |  | 41 (45.1) |  |
| Twenty_four months, n (%) | N | 352 (97.5) | 270<br>(100.0) | 82 (90.1) | <0.001 |
|  | Y | 9 (2.5) |  | 9 (9.9) |  |

(Y:Yes, N:=None)

### 3.2 Correlation between variables in this study

As can be seen in Figure 2, LDH was highly correlated with the ALT variable (coefficient was 0.96), Total Cholesterol was highly correlated with HDL and LDL (coefficients were 0.77 and 0.93, respectively), and NLR was highly correlated with SII (coefficient was 0.9). The above heat map provides the basis for subsequent data downscaling.

**Figure 2.**
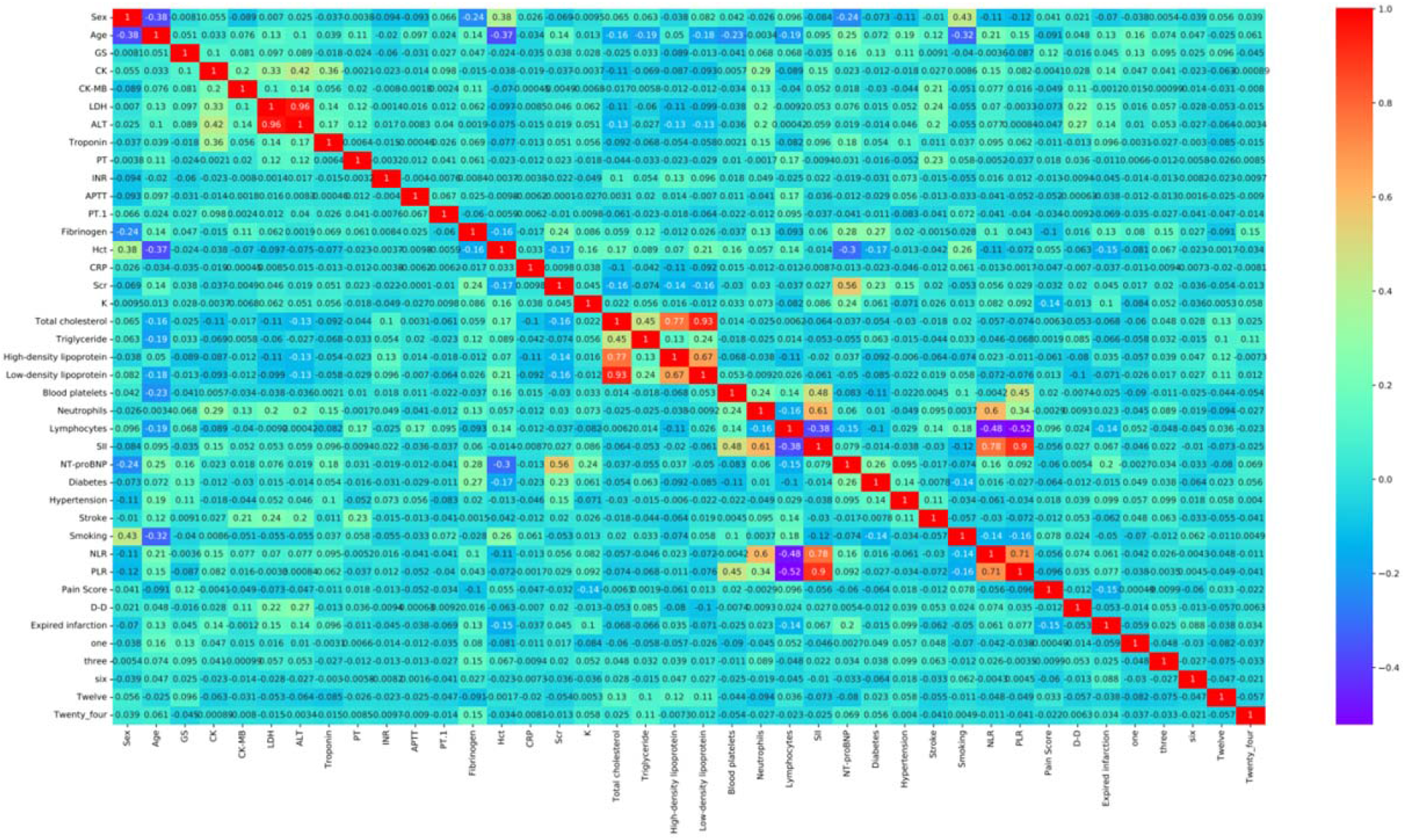
Pearson’s Heat Map.

### 3.3 Development and validation of models

We preprocessed and cleaned the data and randomly divided the dataset into 70% training (n=252) set and 30% testing set (n=109) . Test sets are often representative of a model’s ability to generalize. From Figure 3, Figure 4 and Table 2, it can be seen that the XGBoost model has the best performance. The accuracy was 0.963, the AUC value was 0.97, and the clinical net benefit curves were higher than the remaining two models.

**Table 2.**
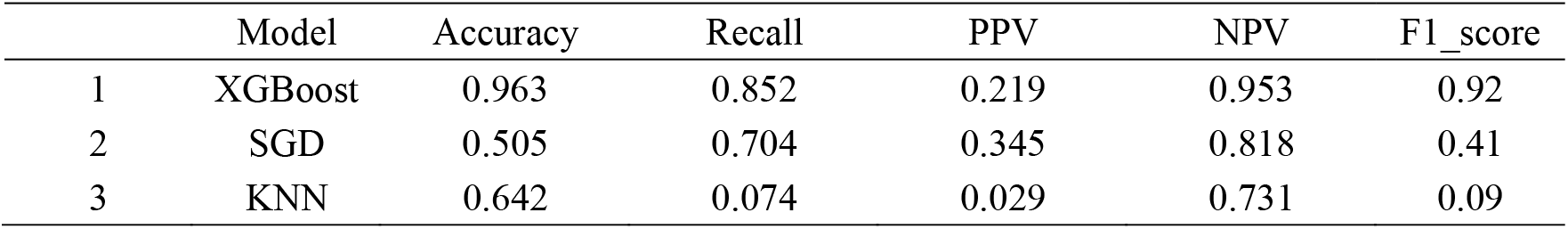
Performance of each model for prediction.

**Figure 3.**
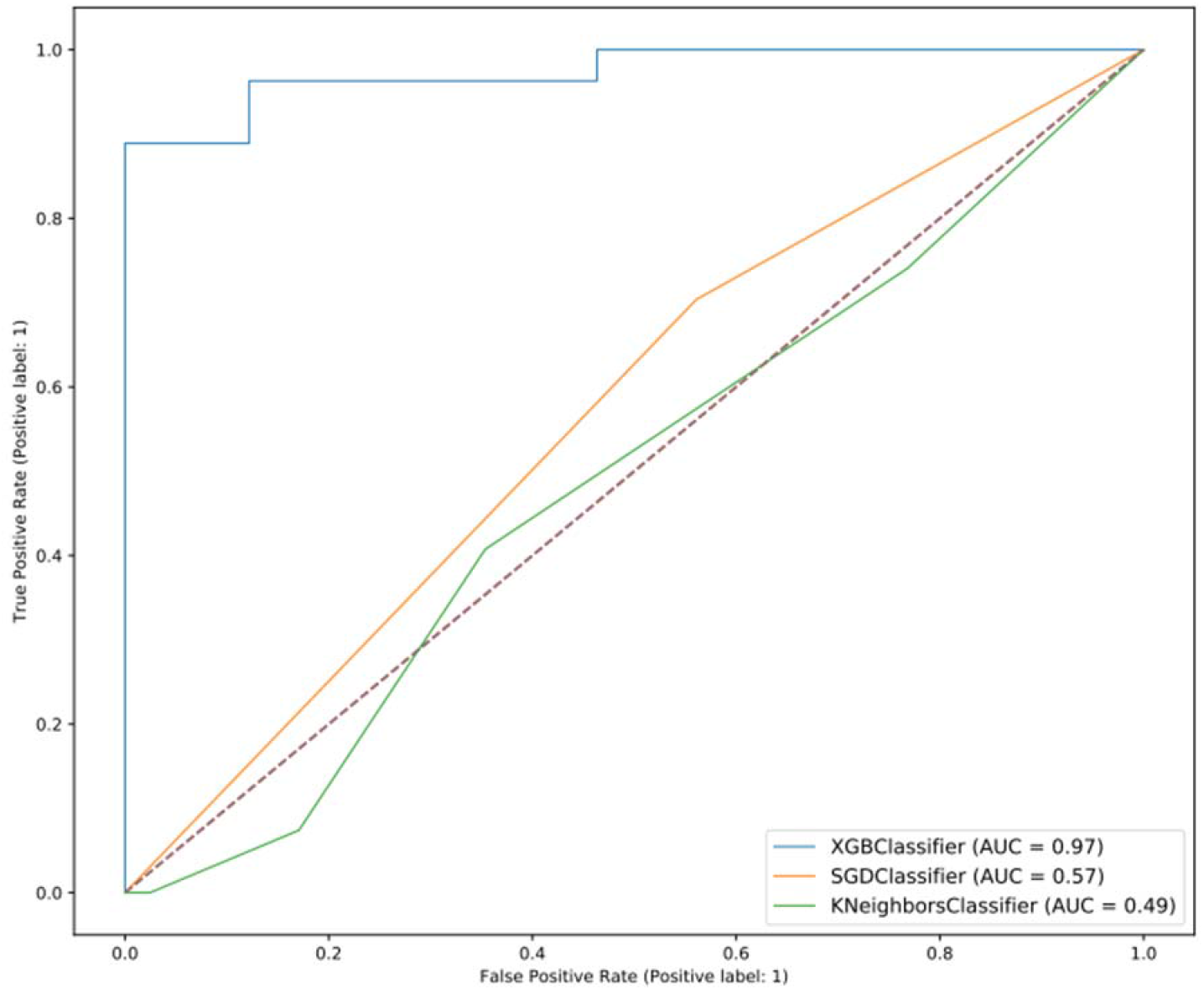
ROC curves for each model.

**Figure 4.**
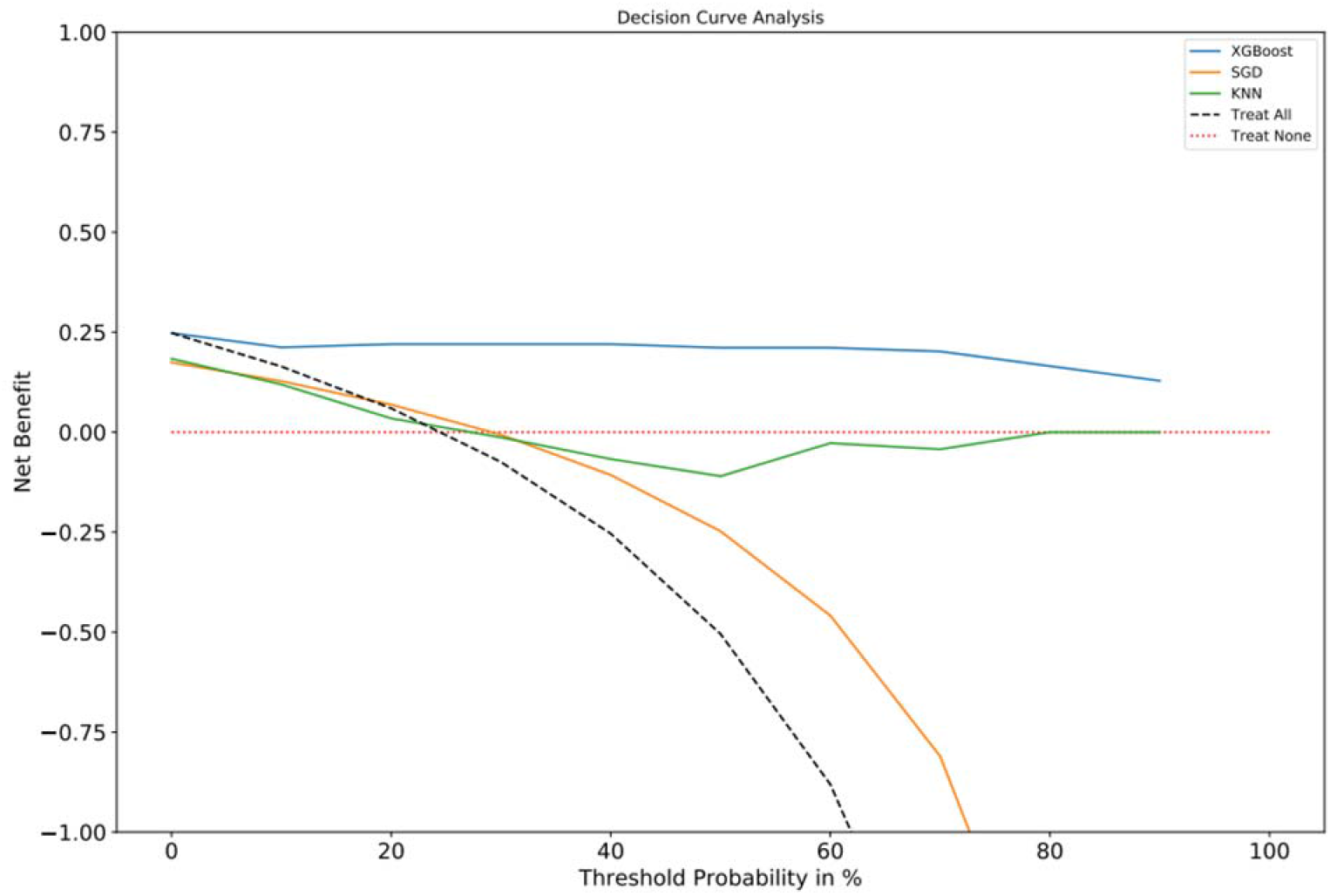
DCA curves for each model.

### 3.4 Observation of feature weights

As can be seen in Figure 5, the weight share of GS is the most important in the whole model. This provides a theoretical basis for selecting variables for our subsequent modeling.

**Figure 5.**
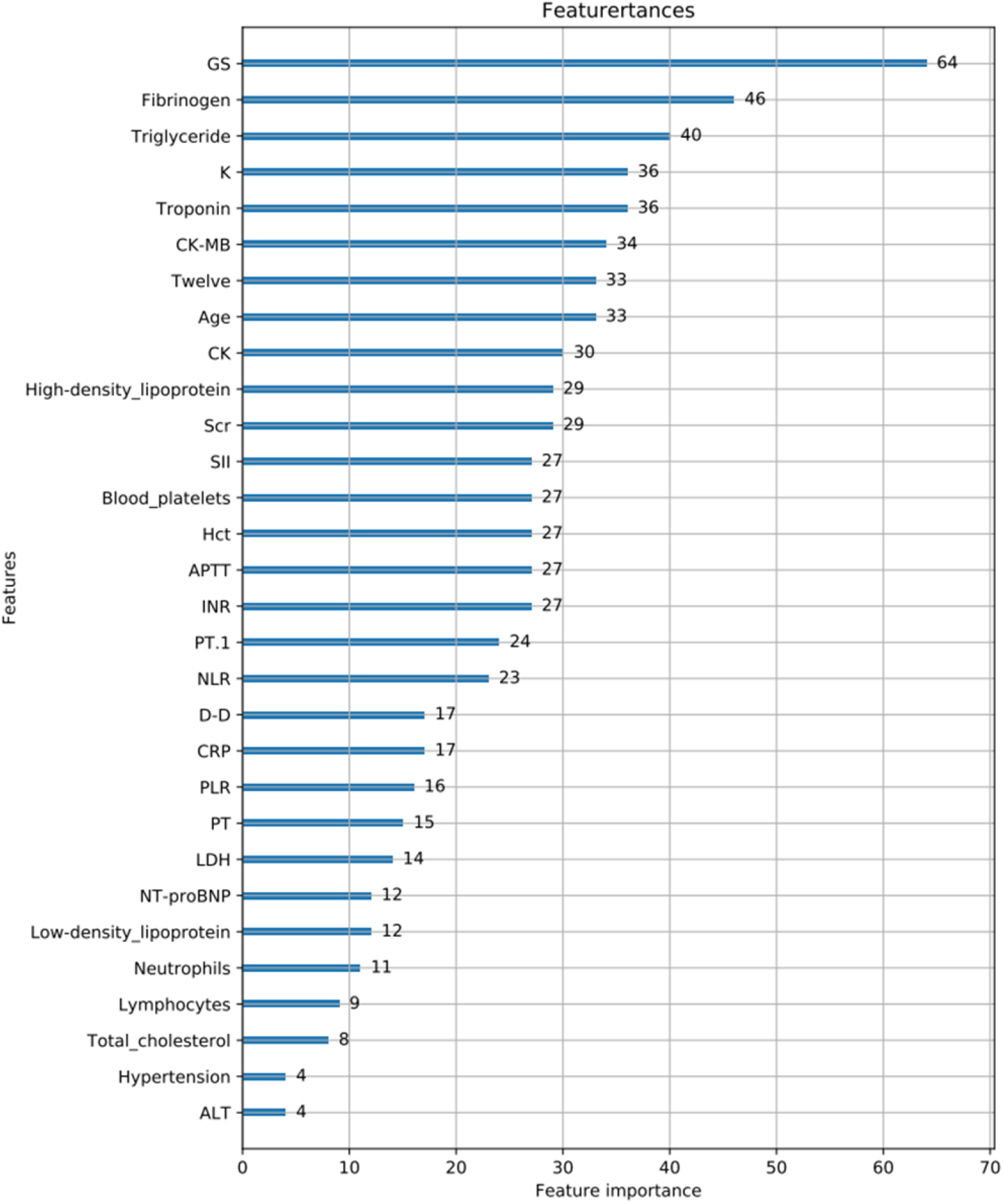
Weighted feature map for XGBoost.

**Figure 6.**
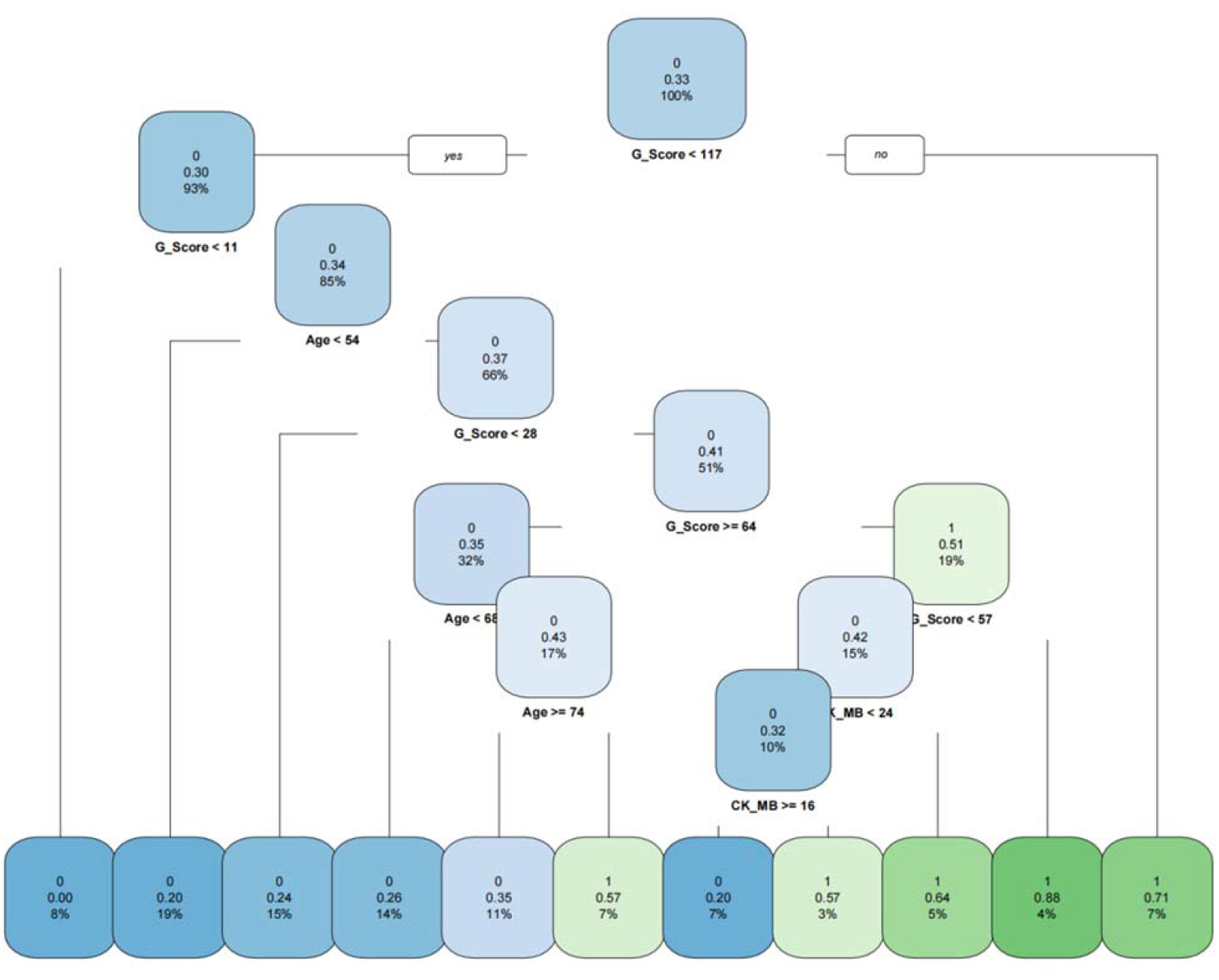
The Decision Tree Model.

### 3.5 Decision Tree Modeling Predicts Likelihood of MACCEs in 2 Years

Based on the weighting characteristics of the XGBoost screening variables and combining the clinical significance, we selected the following variables for model construction: GS, Age, and CK-MB. The Decision Tree (DT) represents a mapping relationship between object attributes and values, so its Cut-off value for each node contributes to the grading of the condition’s hazard level. We assessed the predictive efficacy of the DT by the ROC curve, which has a considerable AUC value of 0.771. In Figure 8, we have utilized the Cut-off values of the decision tree to plot the survival curves with respect to GS.

**Figure 7.**
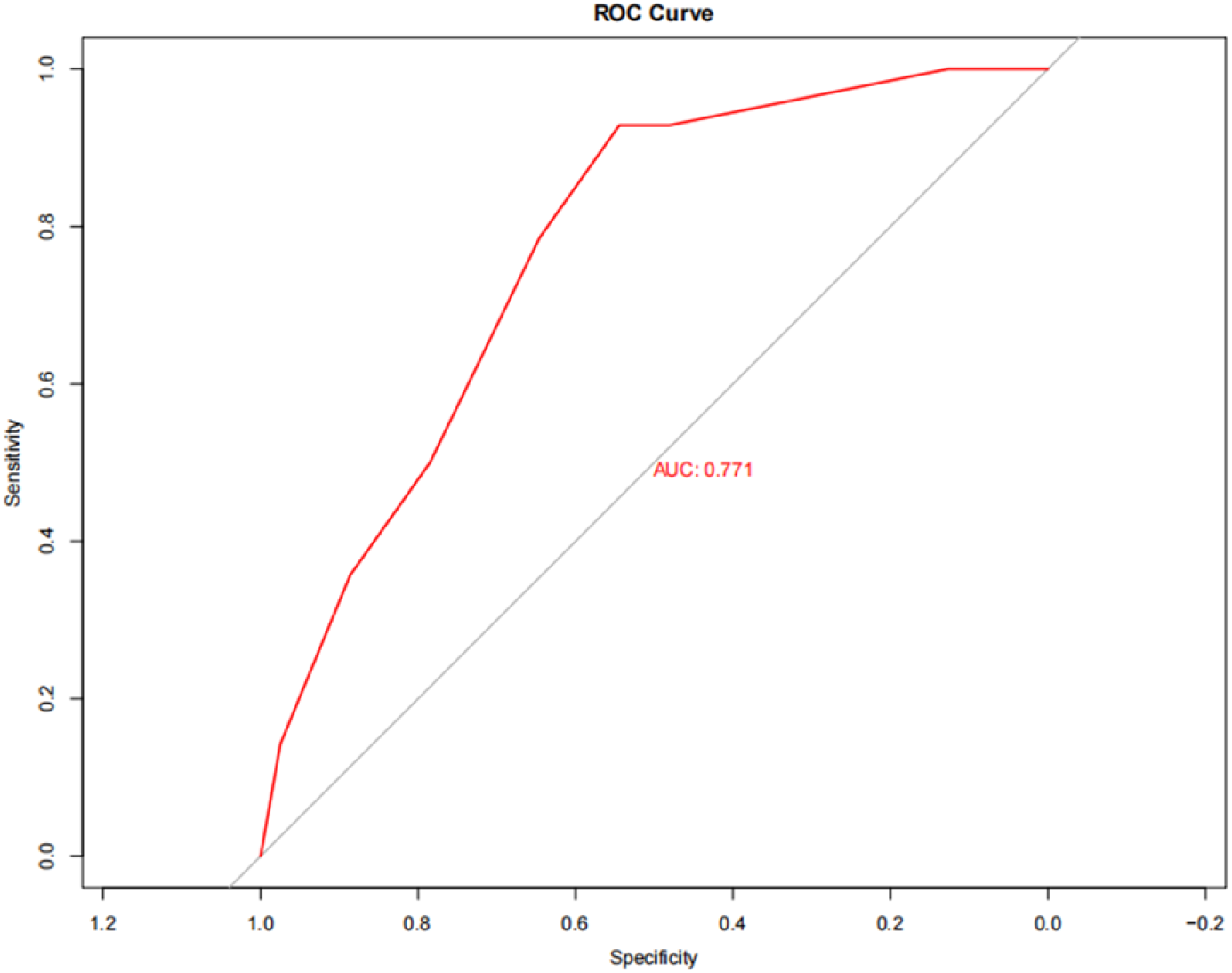
The ROC for Decision Tree Model.

**Figure 8.**
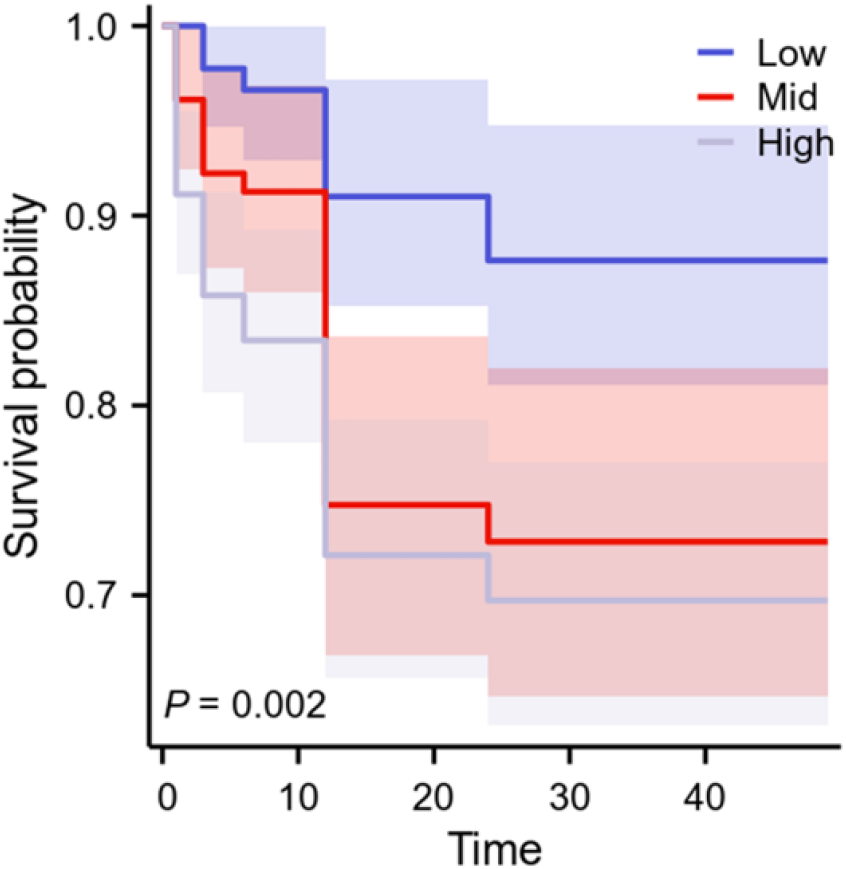
Survival curves for Gensini Scores.

## 4. Discussions

This study utilizes advanced ML methods, precise criteria were applied to the collected clinical characterization variables. Takes advantage of the fact that ML is good at capturing the characteristics of non-linear data. The main features of XGBoost are its simplicity, robustness, and the ability to automatically fill in missing values [22]. It tends to have superior predictive performance in datasets with small samples. While the inner workings of ML remain elusive to us, the Features graph can still help us reveal the characteristics of the data variables. The graph of the weighted characteristics of the variables revealed that GS is a very valuable indicator in predicting the prognosis of ACS patients. The results of this study coincide with the research by Wang et al [14].

The advantage of GS is that it can capitalize on the experience of the coronary operator to quantitatively score the specific degree of stenosis in the coronary artery. Its scoring can be refined down to tiny coronary branches, so it provides a very objective assessment of a person’s plaque load. But this score does not address bifurcation, calcification, and tortuous lesion characteristics. Thus, GS is in a sense more appropriate for patients after emergency PCI where the resident has an approximate estimate of the patient’s prognosis.

However, some of the more established indicators were not remarkable in this study. Examples include SII, NLR, PLR, etc.[11,29]. It may be considered that some patients go directly to the hospital with chest pain and the inflammatory storm in the organism has not yet reached its peak [30,31]. Inflammatory stimuli lead to abnormal activation of platelets, increased thrombus load, and excessive and abnormal endothelial proliferation of blood vessels. The weighted significance of such advanced inflammatory indicators was not found in this small sample study. Perhaps the smaller amount of data caused it.

Acute chest pain of cardiac origin is often characterized by high lethality and their patients themselves tend to have a high number of comorbid clinical conditions. Symptoms such as coronary revascularization, or the occurrence of cerebral infarction or cardiogenic shock are therefore common within a short period of time, so predicting the occurrence of MACCEs is of particular interest. We established this prediction model for this purpose can determine the prognosis of patients more quickly, which can provide a reference for clinical decision-making. In DT, each forked path represents the value of one of the possible attributes, while each leaf node corresponds to the value of the object represented by the path traveled from the root node to that leaf node [28]. And according to the current literature research, there is no relevant study that grades the GS for coronary load with a more rigorous degree of risk. DT fit right in with the original intent of our study.

Based on the Featrues and clinical significance, we selected the above 3 indicators for the following reasons. Kolovou G’research team has demonstrated that age is strongly associated with the development of coronary heart disease [32,33]. Similarly, CK-MB is uniquely specific and sensitive in the diagnosis of ACS [34,35]. Age, CK-MB integrated with GS obtained by coronary angiography results to construct the predictive model. In the DT, the Cut-off value of each node has a more significant result. We can draw on each node value to provide a theoretical basis for grading the severity of coronary heart disease.

And the significance of this prediction model is to actively guide the treatment of patients who do not understand the concept of the condition, to improve their prognosis, and to reduce the incidence of MACCEs within 2 years. Finally, we can see from the survival curves that the lower the GS score, the better the long-term prognosis of the patient, which is consistent with clinical practice.

## 5. Limitations

There are inevitable limitations to this study. First, the study was single-center data. The generalizability of the model is debatable. Second, there is a flaw in the node setting for the follow-up time. It is possible that a model that records the time from admission time to the occurrence of MACCEs would work better. Third, the follow-up studies were all conducted under ideal conditions based on good patient medical compliance and regular medication intake, and did not include other confounding factors after PCI. We are still working on data collection to increase the robustness of subsequent models.

## 6. Conclusions

Our study shows that GS is a persuasive metric for determining the long-term prognosis of ACS patients based on machine learning theoretically investigated. And the DT model constructed using GS, Age and CK-MB indicators had good predictive efficacy for the occurrence of MACCES within 2 years in ACS patients after PCI. The Cut-Off value of GS in the DT can provide a theoretical basis for subsequent grading of coronary risk level.

## Data Availability

The data that support the findings of this study are available on request from the corresponding author upon reasonable request.

## Abbreviations

ACS: Acute Coronary Syndrome
ALT: Alanine Transaminase
APTT: Activated Partial Thromboplastin Time
AUC: Area Under the Curve
CK: Creatine Kinase
CK-MB: Creatine Kinase-Myocardial Band
CRP: C-Reactive Protein
D-D: D-dimer
DT: Decesion Tree
GS: Gensini Score
K: Kalium
KNN: K-Nearest Neighbor
LDH: Lactic Dehydrogenase;
MACCEs: Major Adverse Cardiac and Cerebrovascular Events
ML: Machine Learning
NPV: Negative Predictive Value
NLR: Neutrophil-to-Lymphocyte Ratio
PCI: Percutaneous Coronary Intervention
PLR: Platelet-to-Lymphocyte Ratio
PPV: Positive Predictive Value
PT: Prothrombin time
ROC: Receiver Operating Characteristic Curve
SGD: Stochastic Gradient Descent
SII: Systemic Immune-Inflammation index
TT: Thrombin Time

## 7. Ethics approval and consent to participate

The Ethics Committee of the Fourth Affiliated Hospital of Zhejiang University School of Medicine approved the study.

## 8. Consent for publication

All authors agreed to the publication of the article.

## 10. Authors’ Contributions

Lixia Chen and Sixiang Jia conceived the study. Sixiang Jia completed the dissertation. Sixiang Jia and Xuanting Mou completed the statistical analysis of the data. Yiting Tu reviewed the article and provided suggestions. Lixia Chen, Wenting Lin and Chao Feng were responsible for data organization and collection. Shudong Xia is responsible for the content of the article in its entirety.

## 12. Acknowledgements

Thanks to all the authors for their hard work.

## 13. Availability of data and material

The dataset cannot be shared as it relates to patient privacy.

